# Common genetic modifiers influence cardiomyopathy susceptibility among the carriers of rare pathogenic variants

**DOI:** 10.1101/2024.12.17.24318501

**Authors:** Samantha J. Klasfeld, Katherine A. Knutson, Melissa R. Miller, Eric B. Fauman, Joanne Berghout, Rob Moccia, Hye In Kim

## Abstract

Cardiomyopathy presents significant medical burden due to frequent hospitalizations and invasive interventions. While cardiomyopathy is considered a rare monogenic disorder caused by rare pathogenic variants in a few genes, emerging evidence suggests that common genetic modifiers influence disease penetrance and clinical variability. Quantifying the interplay between common genetic modifiers and rare pathogenic variants is challenging due to the rarity of cardiomyopathy cases and pathogenic variant carriers. In this study, we utilized large-scale genetic and phenotypic data from the UK Biobank to refine the genetic architecture of hypertrophic and dilated cardiomyopathies. Using ClinVar annotations and variant effect prediction tools, we first identified known and predicted pathogenic variants and demonstrated their robust association with disease risk, age of diagnosis, and quantitative cardiac phenotypes that reflect disease progression. We next examined the impact of polygenic risk scores on disease in the combined sets of known and predicted pathogenic variant carriers. Indeed, the polygenic risk scores were significantly associated with increased disease risk, with rare pathogenic variant carriers in the top 20% polygenic risk having 2.6 and 2.4 times higher risk than those in the bottom 20% for hypertrophic and dilated cardiomyopathy, respectively. We observed stronger associations in the carrier sets that included predicted pathogenic variant carriers, suggesting improved statistical power. In summary, our study adds to the evidence that common genetic modifiers influence the cardiomyopathy disease risk among rare pathogenic variant carriers and illustrates the benefit of incorporating variant effect predictions to examine the polygenic influence in rare disease variant carriers.

## Introduction

Cardiomyopathy, a condition affecting the heart muscle, is characterized by impaired cardiac function and is a common cause of heart failure. Cardiomyopathy is classified into subtypes based on distinct abnormalities in cardiac structure and function. Two of the major subtypes are hypertrophic cardiomyopathy (HCM) and dilated cardiomyopathy (DCM). HCM (prevalence: ∼1:543 in the general adult population) is primarily attributed to sarcomere hyperactivity and is characterized by myocardial thickening ^1,2^. On the other hand, DCM (prevalence: ∼1:220) is primarily associated with reduced cardiac contractility and is characterized by dilation of the left ventricle or both ventricles ^2–5^. Approximately 20-50% of cardiomyopathy patients who receive clinical genetic testing carry a rare pathogenic variant in an established cardiac disease gene ^6,7^. These are typically sarcomere and cytoskeletal genes that most commonly follow an autosomal dominant inheritance pattern ^8,9^. Nevertheless, carriers of these variants display low aggregate penetrance of adverse cardiomyopathy ^10^, suggesting that a simple monogenic disease model is insufficient to explain the clinical variability in carriers of these known rare pathogenic variants ^11–13^.

Potential explanations for this variability include differences in lifestyle and traditional cardiovascular risk factors, varying pathogenicity among the causal variants, and differences in disease predisposition attributed to the common genetic background. For the latter, inherited disease risk can be modeled based on the threshold liability framework (Figure S1) ^14^. In this model, an individual who carries a rare pathogenic variant will see their risk shift towards or away from clinically presenting disease based, in part, on the aggregated effect of common genetic modifiers ^14^. While previous studies have shown that common genetic modifiers influence cardiomyopathy disease risk in the general population and in the carriers of rare pathogenic variants ^15–21^, statistical power has been limited due to the rarity of the pathogenic variants and the small number of individuals with cardiomyopathy.

Large-scale genome-or exome-sequencing is essential to ascertain a sufficiently powered cohort of pathogenic variant carriers to examine the effect of common genetic modifiers on disease outcomes among this population. Biobanks, such as the UK Biobank, offer a potential solution with their large sequence data that can be linked to extensive phenotypic information. However, a significant challenge lies in the limited understanding of the functional and clinical effects of the identified variants from these sequencing studies. To this end, computational algorithms that predict the pathogenicity of protein coding variants can be leveraged. AlphaMissense is one of the state-of-the-art prediction algorithms that is based on evolutionary conservation, sequence context, and protein structure, and has demonstrated exceptional performance across several clinical and experimental benchmarks ^22^.

The aim of the study is to understand the effect of common genetic modifiers on disease burden among rare pathogenic variant carriers utilizing the genetic and phenotypic data from 379,721 UK Biobank European participants. We also evaluate whether the use of computational variant effect predictions can increase the pool of pathogenic variant carriers and lead to improved statistical power to detect the effect of common genetic modifiers. Results from this study can contribute to a more refined understanding of the genetic architecture of cardiomyopathy and can offer a generalizable approach to increase statistical power to examine the interplay between common genetic modifiers and rare pathogenic variants.

## Methods

### Subjects and Data

Analyses were conducted using the genetic and phenotypic information from the individuals of self-reported European ancestry (field 22006) in the UK Biobank. All participants of the UK Biobank consented to the use of their genetic and medical information for research purposes. The study was approved by the North West Multi-centre for Research Ethics Committee as a Research Tissue Bank approval. HCM and DCM disease status, select comorbidities and other covariates were extracted from inpatient hospital data, medication data, and self-report data (Table S1). Cardiac size and function measurements were available in the subset of the participants who underwent cardiac MRI imaging ^23^. A total of 2,532 cardiomyopathy cases were identified, including 390 HCM and 1,085 DCM cases (Table S2). Age of diagnosis for HCM and DCM were derived based on the age of diagnosis for cardiomyopathy (field 131338) and the case status for HCM and DCM, respectively. Exome genotype data (n=379,513) were used to derive rare pathogenic variant carriers and imputed genotype data (n=378,606) were used for PRS calculation. Details of the exome sequence and imputed genotype data used in this study are described elsewhere ^24,25^.

### Classification of rare pathogenic variants from exome sequences

Eighteen genes were previously linked to hypertrophic and dilated cardiomyopathy based on the expert-curated evidence according to the ClinGen gene-disease validity framework (Figure S2) ^8,9^. Protein coding variants in the MANE (Matched Annotation from NCBI and EMBL-EBI) transcripts ^26^ were considered. For *TTN*, only the variants within the exons abundantly expressed in heart tissue (percent splice in scores ≥ 90%) ^27^ were considered. Variants were classified as known if they are annotated as pathogenic/likely pathogenic variants (PLP) in ClinVar (accessed in March 2024) [15]. Except for the variants annotated as benign in ClinVar, all other variants were examined by the workflow described in the results and Figure 1A to identify additional likely pathogenic variants. Note that *TTN* was given special handling based on the insights from prior publications and only predicted loss-of-function (pLoF) variants were included.

**Figure 1.**
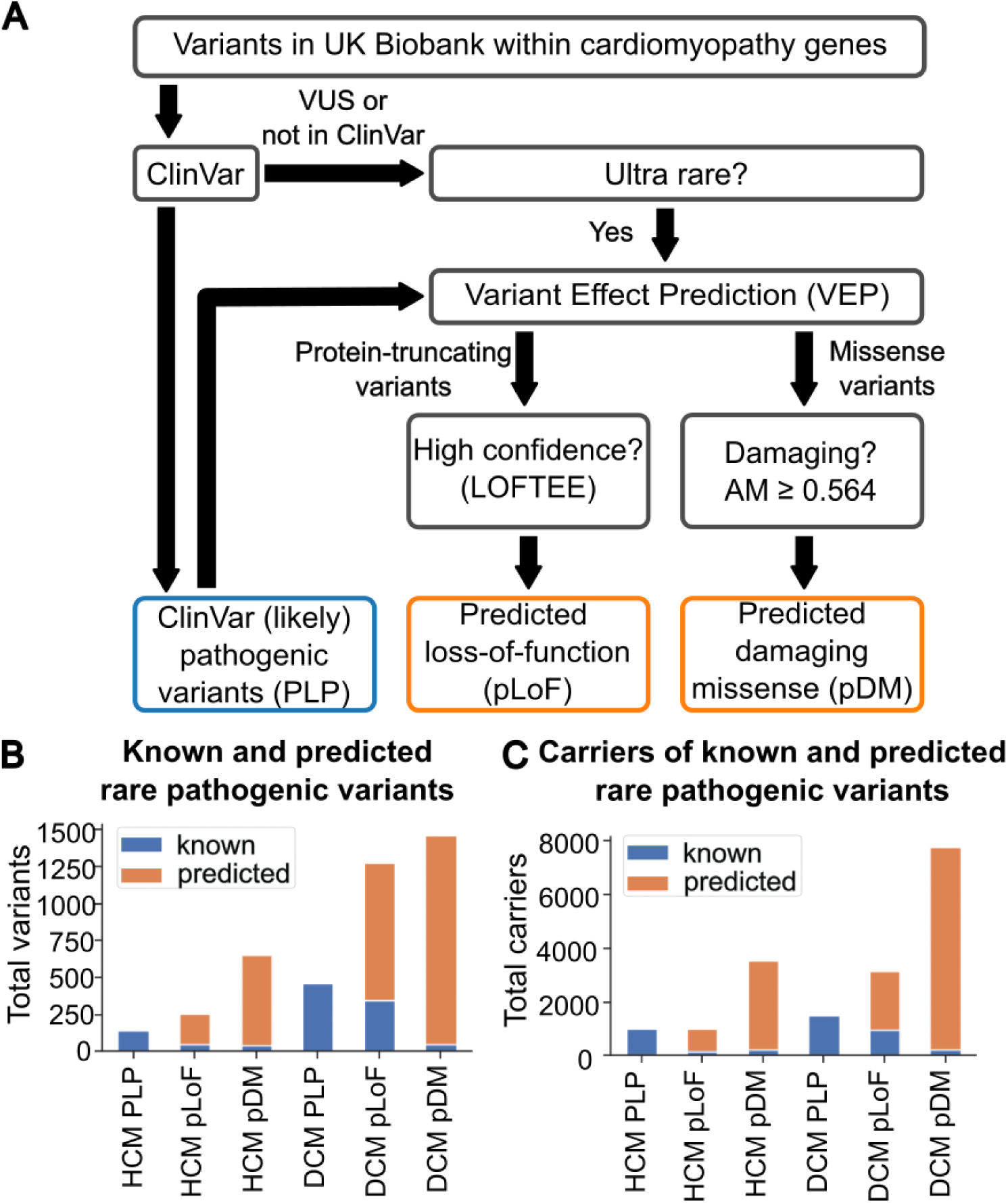
Identification of known and predicted rare pathogenic variants for HCM and DCM in the UK Biobank exome data. (A) Computational workflow used to identify known pathogenic or likely pathogenic (PLP) variants in ClinVar as well as predicted loss-of-function (pLoF) and predicted damaging missense (pDM) variants. (B) Number of known (blue) and predicted pathogenic (orange) variants. (C) Number of carriers of known (blue) and predicted (orange) pathogenic variants. AM, AlphaMissense prediction score.

### Calculation of polygenic risks scores

The list of common genetic variants and their effect sizes were derived from prior genome-wide association studies (GWAS). The HCM PRS was computed from 29 independent variants with significant association (<1% FDR) in the latest HCM GWAS meta-analysis (Table S3) ^28^. We specifically leveraged the sarcomere-negative GWAS (3,860 cases and 38,942 controls) to avoid the inclusion of common variants that may simply tag the effect of rare pathogenic variants ^16,19^. For DCM, due to the scarcity of DCM GWAS and DCM-associated variants available at the time, we instead utilized the GWAS of left ventricular ejection fraction (LVEF), a key diagnostic measure of DCM, in the subset (n=36,041) of the UK Biobank [17] as the substrate for PRS. PRS constructed using LVEF GWAS was strongly associated with DCM status, justifying its use as a substitute for PRS derived from DCM GWAS ^17^. Twenty-two lead variants of the significant signals (P-value < 5e-8) from the LVEF GWAS (Table S4) were used to calculate PRS excluding the subset included in the LVEF GWAS. Both HCM and DCM PRS scores were computed as the total sum of the allelic dosage weighted by the effect size of each variant using the linear scoring function in PLINK2 ^29^. Individuals were categorized into three PRS bins: low (bottom 20%), intermediate (middle 60%), high (top 20%).

### Statistical tests

We examined the contribution of rare pathogenic variants and polygenic risk scores on three disease endpoints: risk, onset, and progression. All models included age, sex, BMI, exome batch, array batch, 10 ancestry principal components, and smoking, diabetes, hypertension, and hypercholesterolemia status as covariates. Age was omitted in the model for disease onset. Cox proportional hazards (CoxPH) models were used to test the association of genetic variants with disease risk. Censor age was derived from the latest date documented in the Electronic Health Record (date at loss to follow-up [191], death from death registry [40000], source censor [54], or last assessment [53]). For all CoxPH models, we report the adjusted hazard ratio and p-value for the variant carrier or PRS term and evaluated significance using a Bonferroni corrected threshold based on the total number of tests.

The CoxPH model assumes that the hazard ratio remains constant, or *proportional*, over time. For each CoxPH model, we tested whether the proportional hazards assumption was met using Schoenfeld residual tests using the ’coxzph’ function in R ^30^. There was a minor violation of the assumption (coxzph p < 0.05) in the models testing the association between pLoF and predicted damaging missense (pDM) pathogenic variants and disease (**Error! Reference source not found.**). To address this, we ran additional CoxPH models adding a time-varying component and confirmed that the hazard ratios across the observed diagnosis age ranges were comparable to the hazard ratios from the original models (Table S6). The proportional hazard assumption was held for all CoxPH models testing the association between PRS and disease (Table S7).

Linear regression models were used to evaluate the effects on age of onset and disease progression, where age of diagnosis and select cardiac measurements were used as proxies for age of onset and disease progression, respectively. To identify the most appropriate proxy phenotypes of disease progression for each cardiomyopathy subtype, we examined available cardiac structure and function measurements for their association with disease status (Table S8). LVEF measurements below 10% (n=27) were considered probable errors and removed from the analyses. Mean left ventricle myocardial wall thickness (LVWT) and left ventricular ejection fraction (LVEF) were most strongly associated phenotypes for HCM and DCM, respectively, and were used for downstream analyses (Figure 4A, 4B).

## Results

### Identifying known and predicted rare pathogenic variants of cardiomyopathy in the UK Biobank exomes

In the exome sequence data of the 379,721 individuals of European ancestry, we found a total of 3,258 and 28,746 protein-coding variants in the 8 and 12 genes previously implicated in HCM and DCM, respectively (Figure S2) 8,9. A small subset of the variants (140 for HCM and 461 for DCM) were previously annotated as pathogenic or likely pathogenic in ClinVar (Table S12) and named “PLP” (pathogenic and likely pathogenic) variants. Variants that are not benign or not annotated in ClinVar and rare (allele frequency <2.2e-4, highest frequency observed among the PLP variants) were examined by several variant effect prediction tools to identify variants predicted to be protein-damaging and putatively pathogenic (Figure 1A). First, Variant Effect Predictor (VEP) was used to bin variants by consequence into those predicted to result in protein-truncation (frameshift, stop-gain, and essential splice site) or missense. Protein-truncating variants with high confidence of producing a true loss-of-function effect by LOFTEE 31 were retained and named “pLoF” (predicted loss-of-function) variants (Table S12). For missense variants, those with AlphaMissense prediction scores >=0.564 were retained and named “pDM” (predicted damaging missense) variants (Table S12). This threshold was previously shown to have 90% precision in classifying damaging mutations when compared to ClinVar 22. As a result, in addition to a small subset of the pLoF/pDM variants that are known PLP variants (Figure 1B, 1C, Table S9, S12), we identified additional 206 (HCM) and 928 (DCM) pLoF variants and 607 (HCM) and 1,410 (DCM) pDM variants that have pathogenic potential (Figure 1B; Table S9). This corresponded to 4,148 and 9,716 additional carriers of predicted pathogenic variants for HCM and DCM, respectively (Figure 1C; Table S9), greatly expanding the number of rare pathogenic variant carriers available for downstream analyses. More details of the rare pathogenic variants and carriers can be found in Tables S9-12.

### Validating the effect of the known and predicted rare pathogenic variants on disease risk, age of diagnosis, and progression

To verify that the predicted rare pathogenic variants identified in our workflow do have clinical impact, we examined their association with cardiomyopathy. To increase the power for association testing, variants were collapsed into PLP, pLoF, and pDM variant classes for HCM and DCM. All associations compared carriers of rare pathogenic variants to noncarriers, reflecting the known autosomal dominant nature of the gene-disease relationship.

For HCM, the PLP variant class was significantly associated with increased disease risk with a hazard ratio (HR_PLP_) of 69.4 (p = 2.0e-245, Figure 2A), confirming the profound impact of these well-interpreted pathogenic variants on HCM risk. The pLoF and pDM variant classes were also robustly associated with increased disease risk with relatively smaller, yet still impressive HRs: HR_pLoF_ = 17.9 (p=1.7e-41) and HR_pDM_ = 6.3 (p = 2.8e-21). We then examined the impact of the rare pathogenic variants on the age of HCM diagnosis among the HCM cases. PLP variant carriers had 6.4 years earlier diagnosis (p = 5.4e-6) while pLoF variant carriers had 7.8 years earlier diagnosis (p = 1.0e-3) compared to the noncarriers on average (Figure 3A). To assess the effect of the rare pathogenic variants on disease progression, we examined their association with mean left ventricular wall thickness (LVWT). Only the PLP variant class showed significant association with increased mean LVWT (β_PLP_ = 0.15 mm, p = 9.8 e-3), while no significant association was observed for the pLoF and pDM variant classes (Figure 4A, 4C; Table S13).

**Figure 2.**
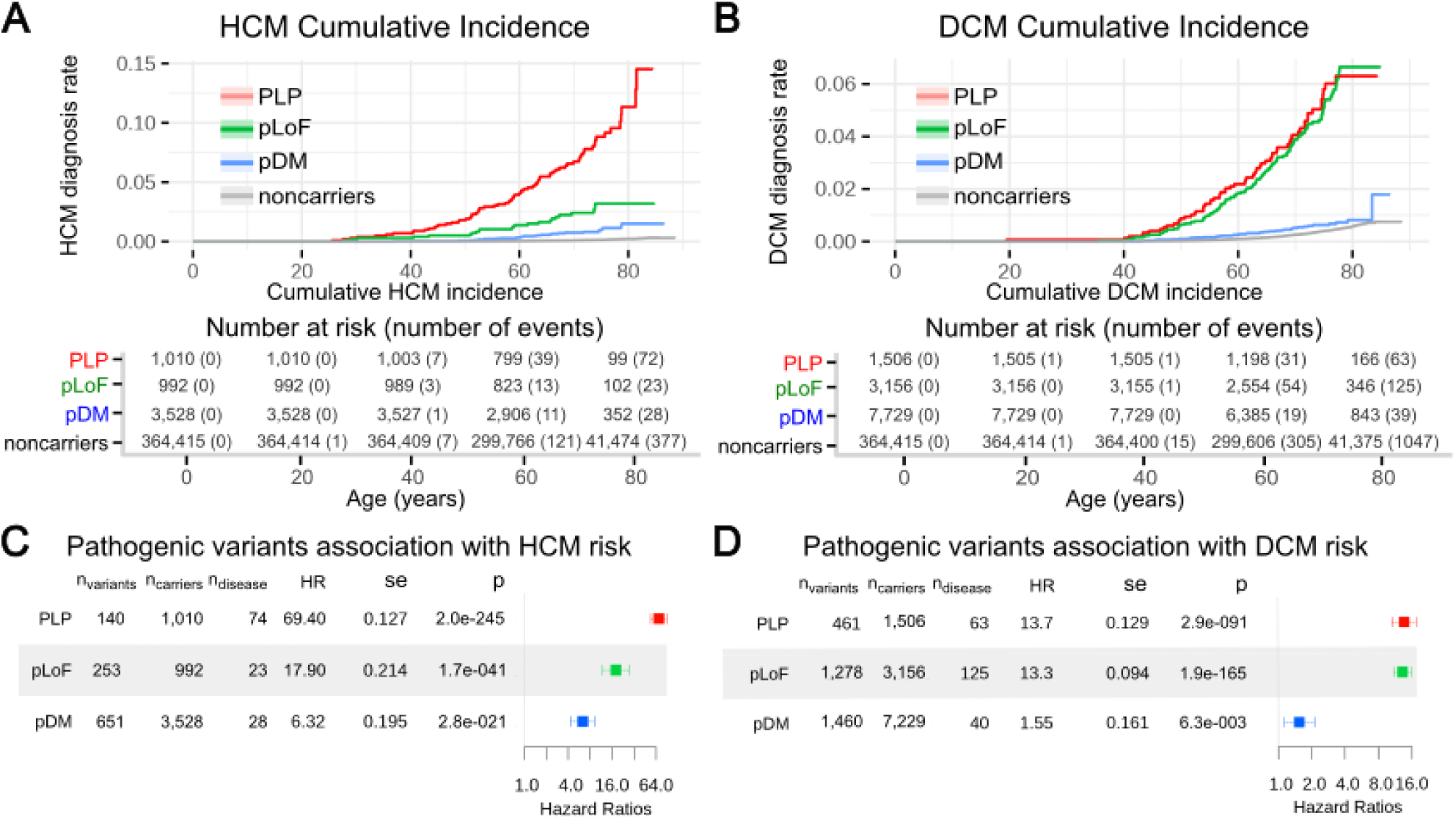
Association of known and predicted rare pathogenic variant classes with HCM and DCM disease risk. Cumulative incidence of HCM or DCM (%) is plotted using Kaplan-Myer (KM) curves (A, B). Forest plots illustrate the association of variant classes with HCM and DCM risk using CoxPH models (C, D). Error bars denote 95% confidence interval. Noncarriers (nc), participants who do not carry known nor predicted rare pathogenic variants; PLP, carriers of known rare pathogenic variants in ClinVar; pLoF, carriers of predicted loss-of-function variants; pDM, carriers of predicted damaging missense variants; n_variants_, number of variants; n_carrier_, number of carriers; n_disease_, number of carriers with disease; HR, hazard ratio; se, standard error; p, p-value.

**Figure 3.**
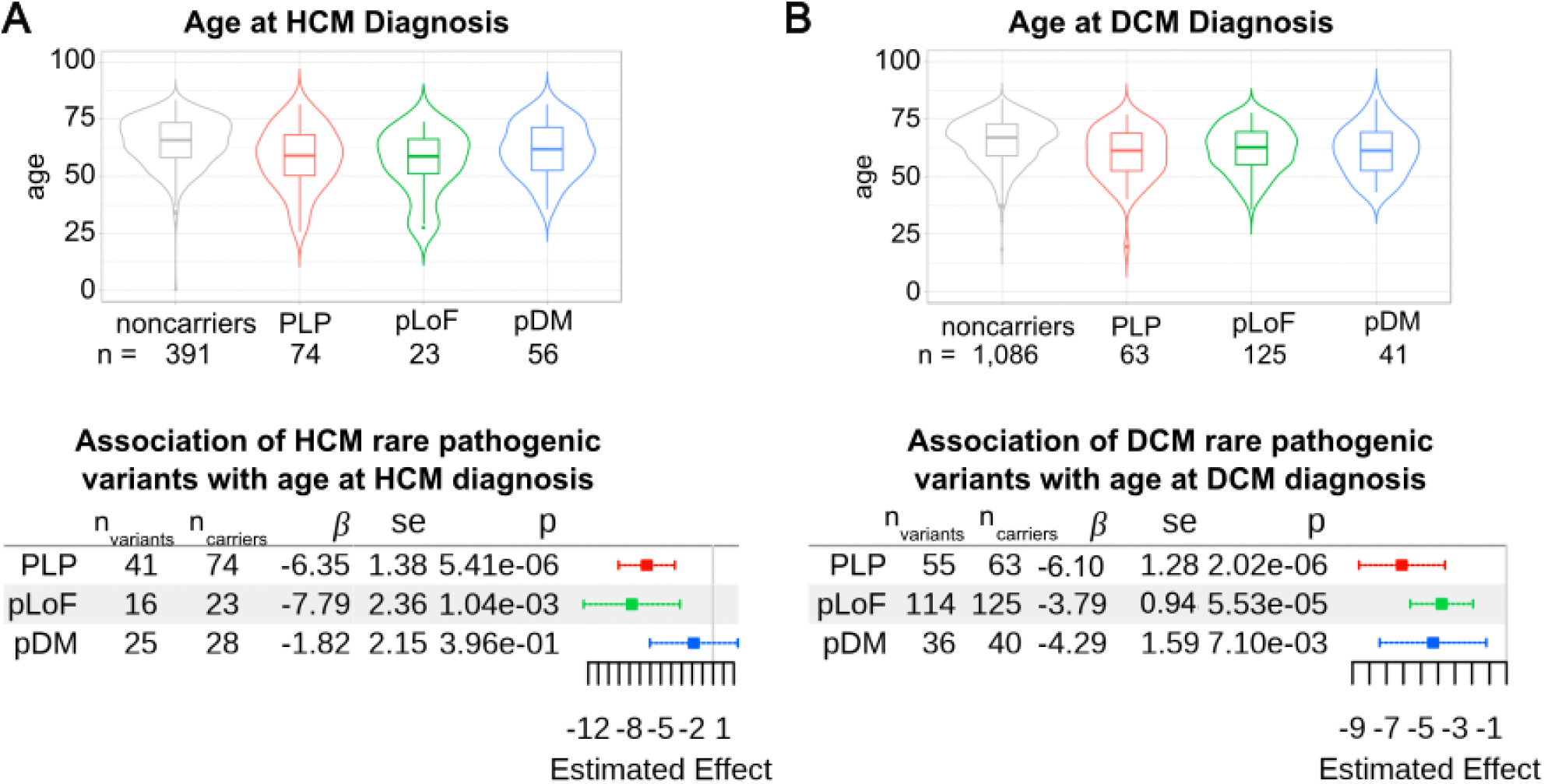
Association of known and predicted rare pathogenic variant classes with HCM and DCM age of diagnosis. Violin and box plots display the distribution of age of diagnosis among HCM (A) and DCM (B) carrier groups (top). Forest plots show the association of rare pathogenic variant classes with age of diagnosis for HCM (A) and DCM (B) using linear regression model (bottom). Error bars denote 95% confidence interval. Noncarriers, participants who do not carry known nor predicted rare pathogenic variants; PLP, carriers of known rare pathogenic variants in ClinVar; pLoF, carriers of predicted loss-of-function variants; pDM, carriers of predicted damaging missense variants; n_variants_, number of variants; n_carrier_, number of carriers; β, estimated effect; se, standard error; p, p-value.

Similarly to HCM, all three DCM variant classes were significantly associated with increased disease risk (Figure 2B). Interestingly, the pLoF variant class exhibited a comparable effect to the PLP variant class (HR_PLP_ = 13.7 [p = 2.9e-91] and HR_pLoF_ = 13.3 [p=1.9e-165]) while the pDM variant class showed a modest effect (HR_pDM_ = 1.6, p = 6.3e-3). All variant classes were significantly associated with earlier age of DCM diagnosis among DCM cases (β_PLP_ = -6.1 yrs, p = 2.0e-6; β_pLoF_ = -3.8 yrs, p = 5.5e-5; β_pDM_ = -4.3 yrs, p = 7.1e-3) (Figure 3B). For DCM, left ventricular ejection fraction (LVEF) was used to assess the effect on disease progression. Both the PLP and pLoF variant classes were also significantly associated with decreased LVEF (β_PLP_ = -4.4 %, β_pLoF_ = -3.8 %), among the DCM cases, but there was no significant association by the pDM variant class (Figure 4B, 4D; Table S14).

**Figure 4.**
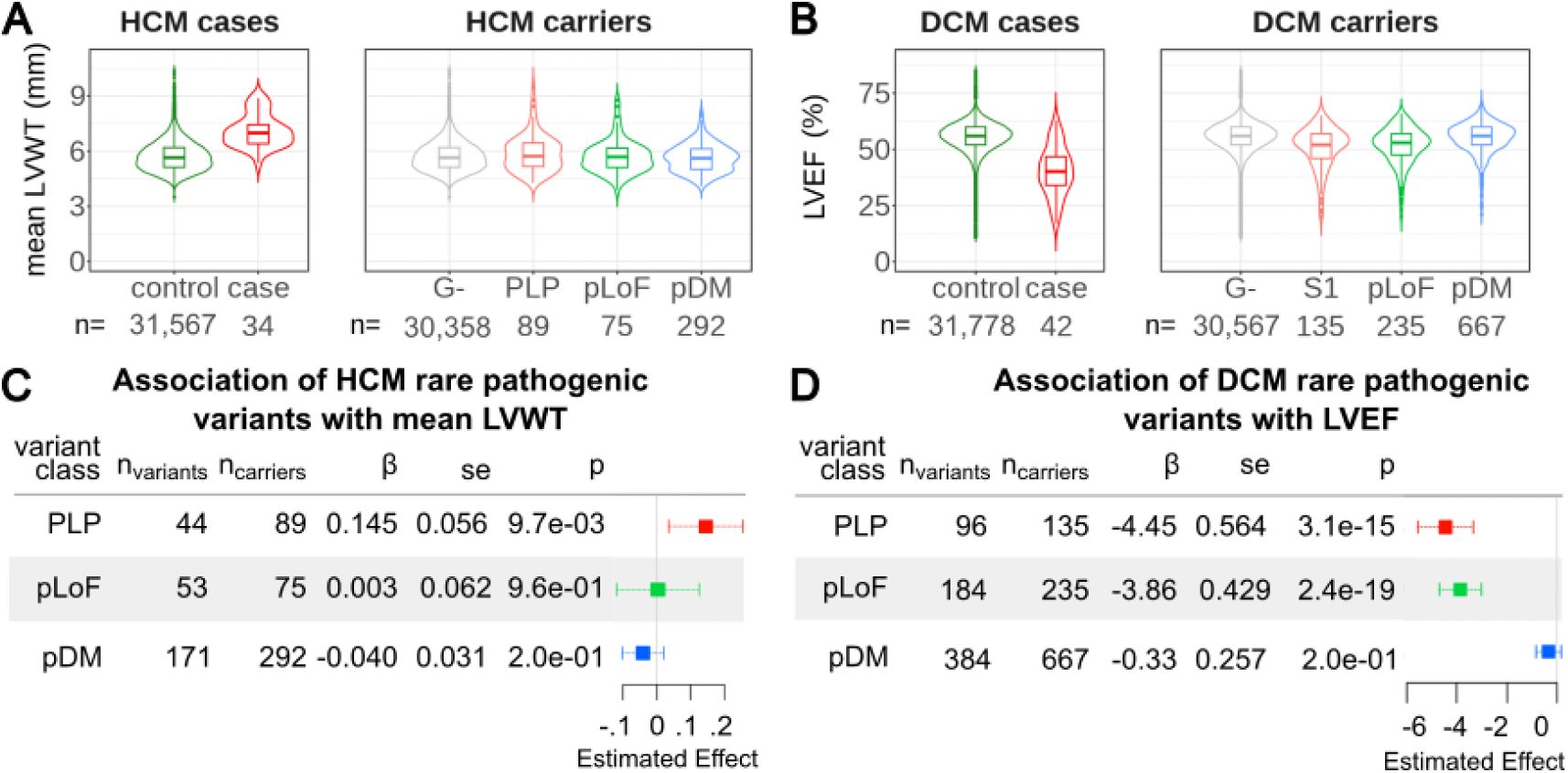
Association of known and predicted rare pathogenic variant classes with cardiac phenotypes representative of HCM and DCM disease progression. (A) Violin plots show the distribution of mean left ventricle myocardial wall thickness (LVWT) per HCM disease status and rare pathogenic variant class (B) Violin plots show the distribution of left ventricle ejection fraction (LVEF) per DCM disease status and DCM rare pathogenic variant class. (C) Association of rare pathogenic HCM variants with mean LVWT using linear regression. (D) Association of rare pathogenic DCM variants with LVEF using linear regression. Error bars denote 95% confidence interval. Noncarriers, participants who do not carry known or predicted rare pathogenic variants; PLP, carriers of known rare pathogenic variants in ClinVar; pLoF, carriers of predicted loss-of-function variants; pDM, carriers of predicted damaging missense variants; n_variants_, number of variants; n_carrier_, number of carriers; β, estimated effect; se, standard error; p, p-value.

### Evaluating the effect of common genetic modifiers on disease risk, age of diagnosis, and progression among the rare pathogenic variant carriers

We next sought to investigate whether common genetic modifiers influence disease in the rare pathogenic variant carriers. To maximize statistical power, we aggregated known and predicted rare pathogenic variant carriers into three carrier sets: set 1 (S1) only includes the PLP carriers; set 2 (S2) includes S1 plus pLoF variant carriers; and set 3 (S3) includes S2 plus pDM variant carriers.

HCM PRS was calculated using the 29 variants identified in the latest HCM GWAS and their reported effects on HCM risk^28^. As expected, the distribution of HCM PRS was right shifted among the HCM cases (median = 0.19) compared to the controls (median = -0.06) (Figure 5A, shown in the S3 carrier set). Carriers and noncarriers in high (top 20%) PRS exhibited higher cumulative incidence of HCM compared to those in low (bottom 20%) PRS (Figure 5B, shown in the S3 carrier set). We confirmed that HCM PRS was significantly associated with increased HCM risk in the noncarrier set (HR_noncarrier_ = 1.89 [p = 2.0e-40]) (Figure 5C). HCM PRS also illustrated a risk-increasing effect in all three carrier sets, but the association reached statistical significance only in the S2 and S3 sets which included predicted rare pathogenic variant carriers (HR_S1_ = 1.25 [p = 5.3e-2], HR_S2_ = 1.48 [p = 6.2e-3], HR_S3_ = 1.45 [p = 1.1e-3]) (Figure 5C).

**Figure 5.**
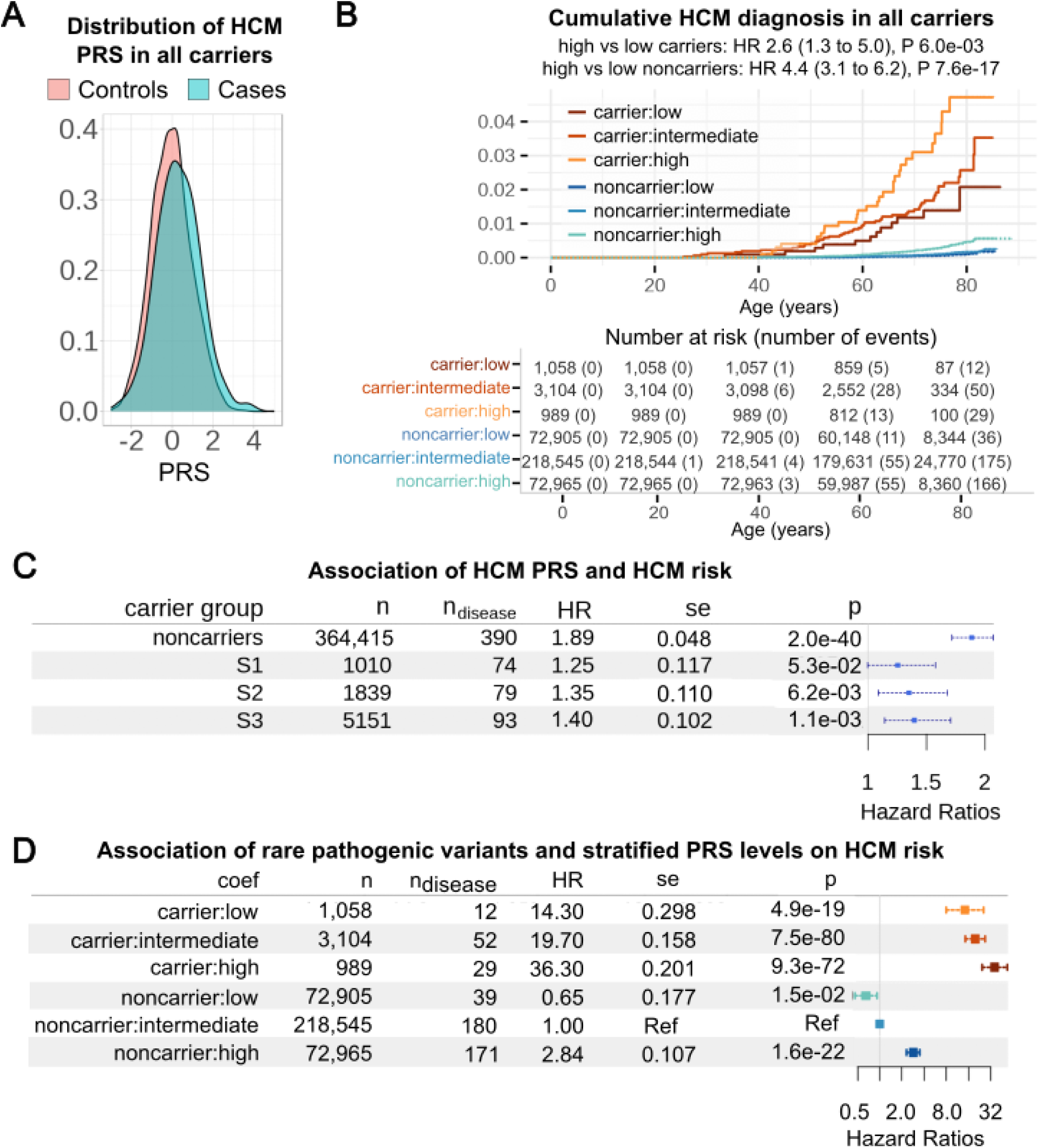
HCM PRS is associated with disease risk among the carriers of known and predicted rare pathogenic variants. HCM PRS distribution is shifted upward in the HCM cases compared to controls among the rare pathogenic carriers (S3) (A). Cumulative incidence of HCM (%) among the rare pathogenic variant carriers (S3) and noncarriers stratified by the HCM PRS is plotted using KM curves with the number at risk and number of events at each timepoint summarized in the table below (B). Forest plot illustrates the association of PRS with HCM risk using CoxPH models among the noncarrier and carrier sets (S1, S2, S3) (C). Forest plot shows the effect of both the rare pathogenic variants and common variant PRS on HCM risk with the noncarriers with intermediate PRS as the reference (D). Error bars denote 95% confidence interval. PRS bins: low (bottom 20%), intermediate (middle 60%), high (top 20%); n, number of individuals in carrier sets; n_disease_, number of HCM cases in the carrier sets; HR, Hazards Ratio; se, standard error; p, p-value.

Notably, the effect of HCM PRS was relatively smaller in the carrier sets compared to the noncarrier set with significant statistical interaction (β_interaction_ = -0.302, p = 7.8e-3 in S3, Table S15). In the S3 set, carriers with high PRS had 2.6-fold higher risk of HCM compared to carriers with low PRS (p = 0.006). Comparing to the noncarriers with intermediate PRS, carriers with high and low PRS had 36.3 and 14.3 higher risk of HCM, respectively (Figure 5D), illustrating a considerable impact of common genetic modifiers on disease risk among the rare pathogenic variant carriers. Contrary to the strong effect of rare pathogenic variants observed, HCM PRS was not associated with age of HCM diagnosis in both noncarrier and carrier sets (Figure S3A, S3B), suggesting a minimal polygenic influence on HCM age of onset. While HCM PRS was significantly associated with mean LVWT, a proxy phenotype for HCM progression, in the noncarriers (β_meanLVWT_ = 0.04 [p = 4.6e-22]), we did not observe significant association in the carrier sets (Figure S3C, S3D) although a nominal significance with a relatively large effect was observed in the S1 set (β_LVEF_ = 0.24 [p = 0.03]).

DCM PRS was computed using the 22 variants identified in the GWAS of LVEF _17_. DCM PRS was right shifted among the DCM cases (median = 0.40) compared to the controls (median = 0.03) (Figure 6A, shown in the S3 carrier set). Carriers and noncarriers with high PRS showed higher cumulative incidence of DCM compared to those with low PRS (Figure 6B, shown in S3). As expected, DCM PRS was significantly associated with higher DCM risk in the noncarrier set (HR_noncarrier_ = 1.46 [p = 1.22e-32]). DCM PRS also showed risk-increasing effect in all three carrier sets, but statistical significance was only met in the S3 set which included the largest number of rare pathogenic variant carriers (HR_S3_ = 1.33 [p = 6.3e-4]) (Figure 6C). In the S3 set, carriers with high PRS had 1.5-fold elevated risk of DCM compared to carriers with low PRS (p=0.002). With noncarriers with intermediate PRS as the reference, carriers with high and low PRS had 7.2 and 3.1 higher risk of DCM, respectively (Figure 6D). While DCM PRS was significantly associated with earlier age of DCM diagnosis in the noncarrier set (β_DCMage_ = -1.07 [p = 5.1e-4]), no effect was observed in the carrier sets (Figure S3E, S3F). As DCM PRS was derived using the LVEF GWAS in the UK Biobank, the effect on DCM progression could not be examined.

**Figure 6.**
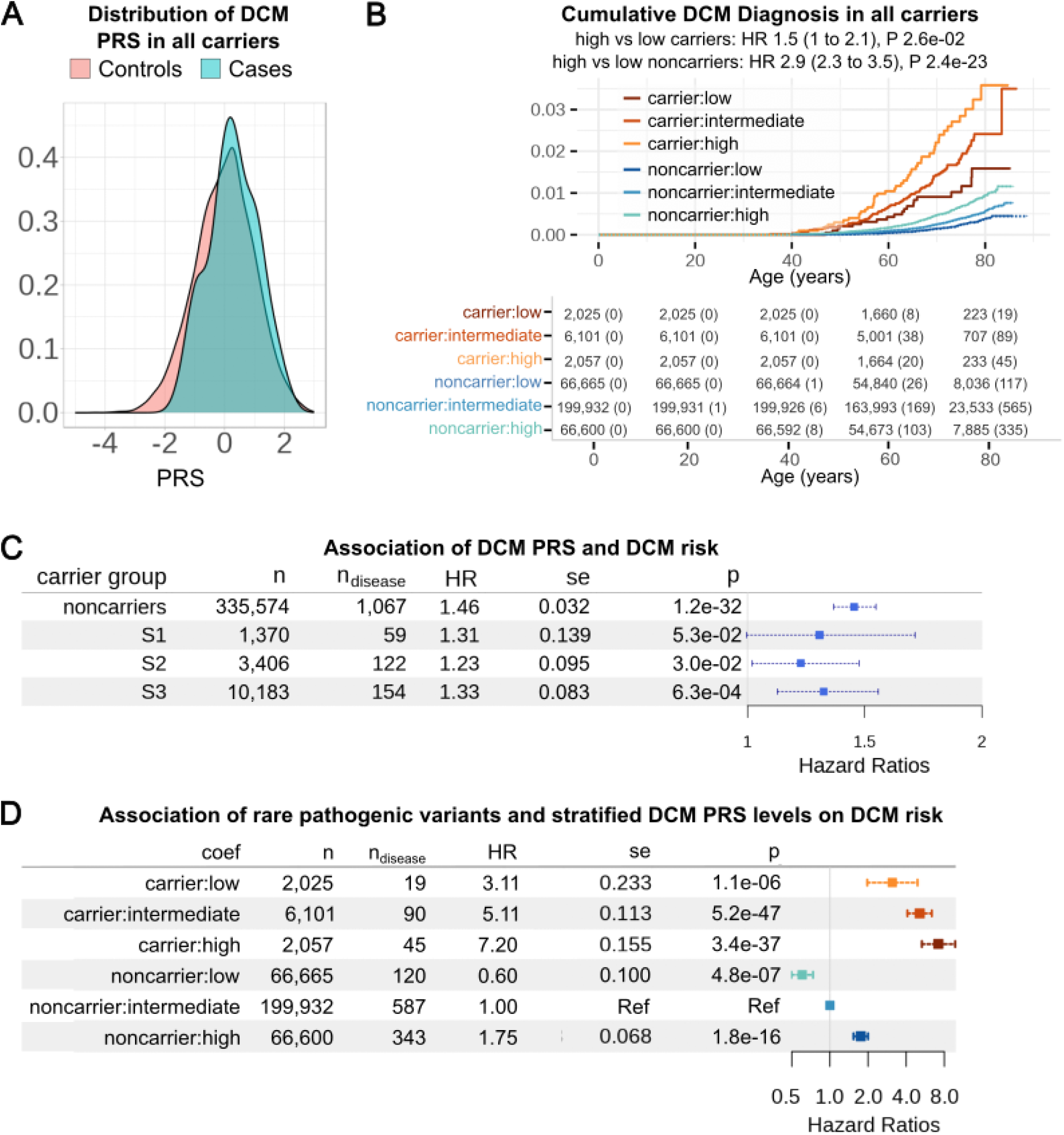
DCM PRS is associated with disease risk among the carriers of known and predicted rare pathogenic variants. DCM PRS distribution is right shifted in the DCM cases compared to controls among the rare pathogenic variant carriers (S3) (A). Cumulative incidence of DCM (%) among the rare pathogenic variant carriers (S3) and noncarriers stratified by DCM PRS is plotted using KM curves with the number at risk and number of events at each timepoint summarized in the table below (B). Forest plot illustrates the association of DCM PRS with DCM risk using CoxPH models among the noncarrier and carrier sets (S1, S2, S3) (C). Forest plot demonstrates the effect of both the rare pathogenic variants and common variant PRS on DCM risk with the noncarriers with intermediate PRS as the reference (D). Error bars denote 95% confidence interval. PRS bins: low (bottom 20%), intermediate (middle 60%), high (top 80%); n, number of individuals in carrier sets; n_disease_, number of DCM cases in the carrier sets; HR, Hazards Ratio; se, standard error; p, p-value.

## Discussion

While cardiomyopathy has been traditionally regarded as a rare monogenic disorder, considerable variability is observed in disease penetrance. While recent studies have suggested that common genetic modifiers alter cardiomyopathy disease risk ^15,16^, examining their effect among rare pathogenic variants has suffered from limited statistical power given the rarity of the cardiomyopathy cases and the pathogenic variants. In this study, we illustrated the benefit of applying computational variant effect predictions to the large exome sequence data from the UK Biobank to expand the pool of rare pathogenic variant carriers. Our results demonstrated that predicted pathogenic variant classes are significantly associated with increased disease burden, suggesting that they are enriched for true pathogenic variants. Increasing the number of rare pathogenic variant carriers by including pLoF/pDM variant carriers in addition to PLP variant carriers strengthened the association observed for the effect of common modifiers on disease risk, indicating improved statistical power. These findings suggest that incorporating variant effect predictions could be a useful and generalizable approach to examine the polygenic influence in other rare diseases.

While we observed significant associations of the predicted pathogenic variants with disease risk in aggregate, it is reasonable to expect that not all predicted variants are truly pathogenic. For example, HCM pLoF and pDM variant classes had weaker association p-value and effect size compared to the PLP variant class (Figure 2). There are two possible explanations for this: 1) the predicted variant classes may include non-pathogenic variants diluting the association or 2) the PLP variants are biased towards variants with stronger effects, i.e., have higher chances of being documented previously. The benefit of incorporating predictions was more apparent for DCM, where pLoF variant class not only had a comparable effect size but also had a stronger p-value compared to the PLP variant class presumably due to the larger number of variants included. This suggests that the pLoF variants in DCM genes are enriched for true high impact variants, warranting further validation and curation of these variants.

In this study, we collapsed known and predicted rare pathogenic variants and carriers across all genes implicated in HCM and DCM to maximize statistical power, but there are inherent limitations in this approach. First, different genes may have different molecular mechanisms, i.e., gain-or loss-of-function, associated with disease. Second, different genes may have different maximal effects on disease so aggregating variants across different genes may introduce noise. Third, genes involved in different biological pathways can have varying degree of influence from the common genetic modifiers. Convergence in the biological pathways influenced by the rare pathogenic variants and common genetic modifiers can in fact offer a possible explanation for the smaller effect of HCM PRS observed among the rare pathogenic variant carriers than among the noncarriers. For instance, if a rare pathogenic variant leads to partial loss of expression or function of a protein or impairs a downstream pathway, a modifier that affects the same protein or pathway will likely have less impact on disease. While it was not feasible with currently available data, larger genetic data and improved understanding of the molecular mechanisms of the genes can enable more accurate assessment of the impact of common genetic modifiers at the individual gene level and provide a more complete picture of the interplay between common genetic modifiers and rare pathogenic mutations.

While phenotypic data from the UK Biobank provided valuable substrates, there are also several limitations that need to be considered. As the UK biobank recruitment was based on voluntary participation, it is noted that it is biased towards healthy individuals ^32^ and may miss individuals with severe rare diseases. In addition, disease case status is mainly derived from ICD codes which are primarily used for billing purposes and may not accurately capture the true disease status, e.g., missed or erroneous codes. Reflecting these limitations, we observed much lower prevalence and penetrance of cardiomyopathy in the UK Biobank [HCM: 0.11%, DCM: 0.30%] compared to the estimates reported in other studies [HCM: 0.15%-.23%, DCM: 0.39%-.53%] ^2^. This bias may have influenced the effect estimates of the rare pathogenic variants and common modifiers on cardiomyopathy risk reported in this study. In addition, our study focused on the European ancestry, which constitutes the majority of the UK Biobank, was adequately powered for statistical analyses, and had prior GWAS available for PRS calculation. It is important to note that the magnitude of effects may differ depending on ancestral backgrounds and future studies using more diverse genetic datasets could provide more accurate effect estimates in other ancestral populations.

In addition to the effects on disease risk, this study also examined the effects of rare pathogenic and common modifier variants on disease onset and progression using age of diagnosis and select cardiac phenotypes (mean LVWT for HCM and LVEF for DCM). Despite the limited number of cardiomyopathy cases in the UK Biobank, we found significant association of rare pathogenic variants with age of diagnosis for both HCM and DCM, demonstrating the effect of rare pathogenic variants on disease onset. Interestingly, HCM PRS did not have an apparent effect on age of diagnosis while DCM PRS was significantly associated with reduced age of diagnosis (Figure S3, B and F). While this could be due to the difference in the number of HCM and DCM cases, it suggests a potential difference in the polygenic effect on age of onset between the two cardiomyopathy subtypes. While the association did not reach statistical significance after adjusting for the number of tests performed, HCM PRS was nominally associated with mean LVWT with a relatively large effect in the S1 carrier set (Figure S3D). It would be of interest to replicate this observation with maximum LVWT ^33^, a more suitable biomarker of HCM than mean LVWT, once publicly released and with a larger number of measurements that will become available as the UK Biobank imaging study further progresses ^23^.

In conclusion, our study highlights the important contribution of common genetic modifiers in determining the inherent risk of HCM and DCM in individuals carrying rare pathogenic variants. This finding offers explanation for part of the observed variability in clinical presentations and suggests potential value of utilizing polygenic risk scores in further stratifying the risk among the rare pathogenic variant carriers. This approach could enable more precise and personalized risk predictions and early diagnosis and intervention for individuals at high-risk and has the potential to enhance patient outcomes and alleviate the overall disease burden of cardiomyopathy.

## Data Availability

The underlying genetic and phenotypic data used in this work are available from the UK Biobank. All data produced in this study are either provided in the article and supplemental information or are available upon reasonable request to the corresponding authors.

## Data availability

The underlying genetic and phenotypic data used in this work are available from the UK Biobank. GWAS summary statistics used to generate polygenic risk scores are available from prior publications ^17,28^. All other relevant data are provided in the article and supplemental information. Any other data or codes may be available upon reasonable request to the corresponding authors.

## Acknowledgements

We thank the participants and research staff of the UK Biobank study. This work was conducted under UK Biobank application 26041. We thank the Postdoc Program of Pfizer Research and Development for their support and guidance for S.J.K.

## Author Contributions

S.J.K. performed the analyses. K.A.K. advised statistical analyses. H.K. and R.M. supervised the work. J.B., R.M., and H.K. conceived the study. S.J.K. and H.K. wrote the initial draft. All authors contributed to the analysis design and review and revision of the writing.

## Supplemental Figures

**Figure S1.**
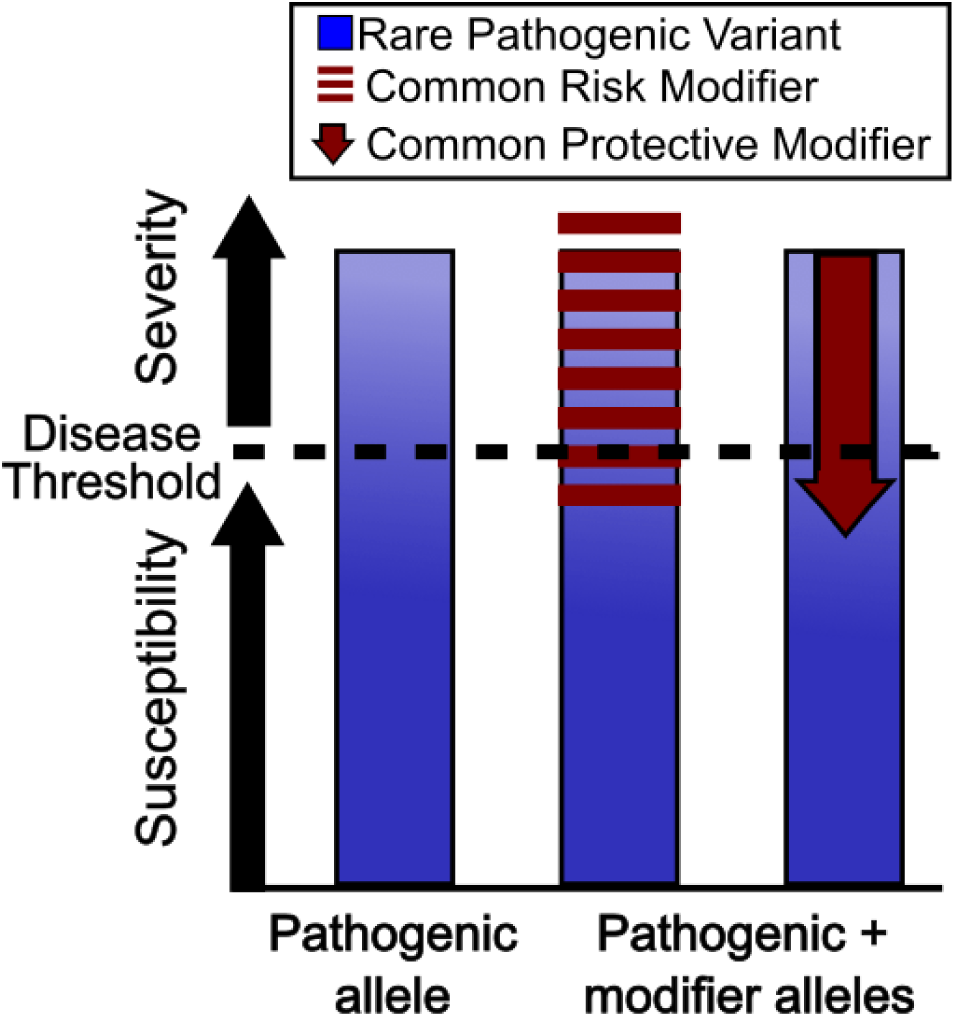
Inherited threshold model of disease ^14^. Rare pathogenic variants (blue) have large influence on disease susceptibility and severity. In a pure monogenic disease model, these variants are fully sufficient to cause disease (left-most bar). However, the monogenic model does not explain the observed variability in disease penetrance and severity. Alternatively, common genetic modifiers may increase (middle bar) or decrease (right-most bar) disease susceptibility and severity in the presence of rare pathogenic variants. Not shown are environmental modifiers that can also affect disease risk and severity.

**Figure S2.**
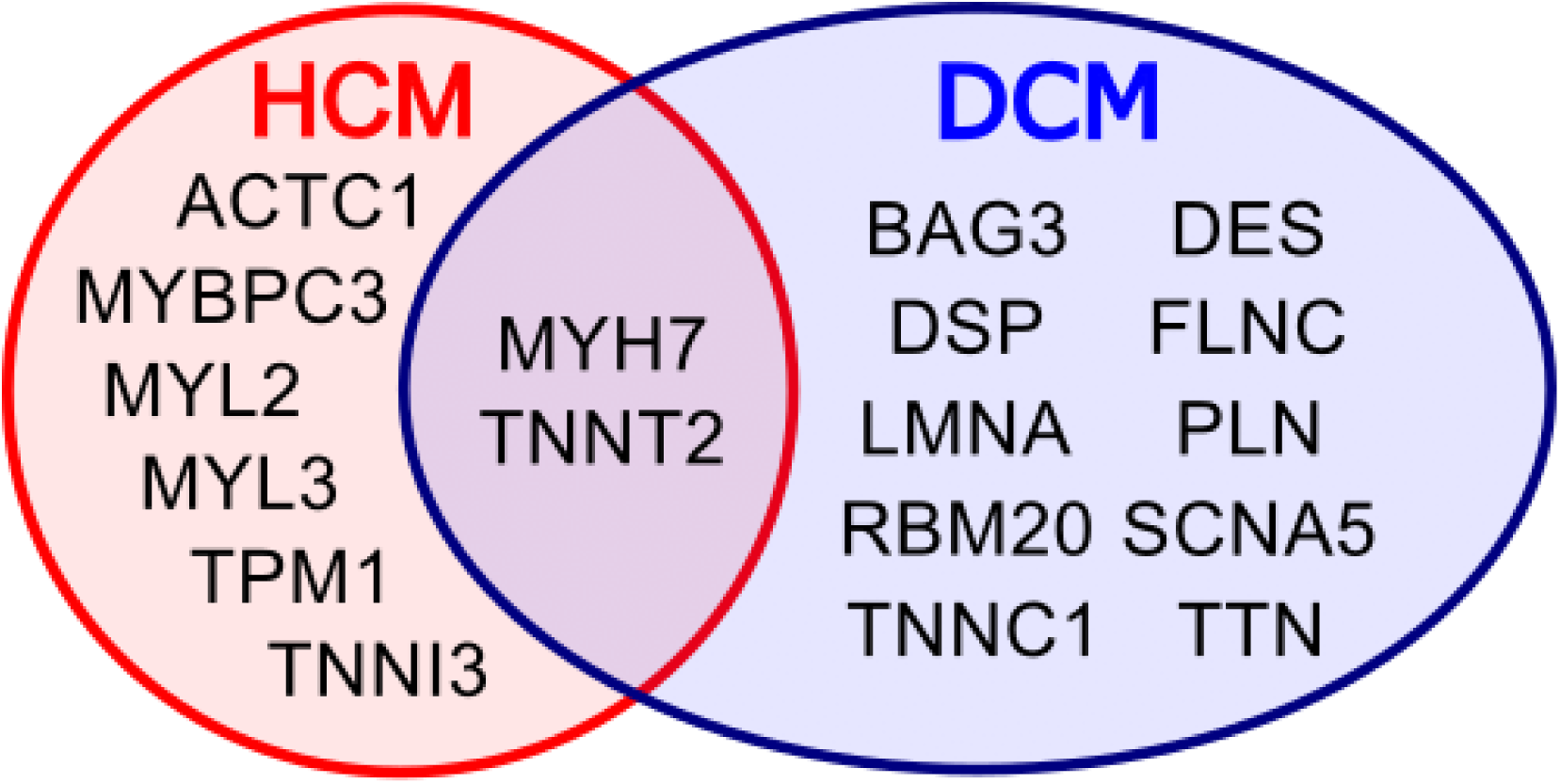
Eighteen genes causally implicated in dilated cardiomyopathy (DCM) and hypertrophic cardiomyopathy (HCM) based on curated evidence classifying the gene-disease relationship as “strong” or “definitive” according to the ClinGen framework^8,9,34,35^.

**Figure S3.**
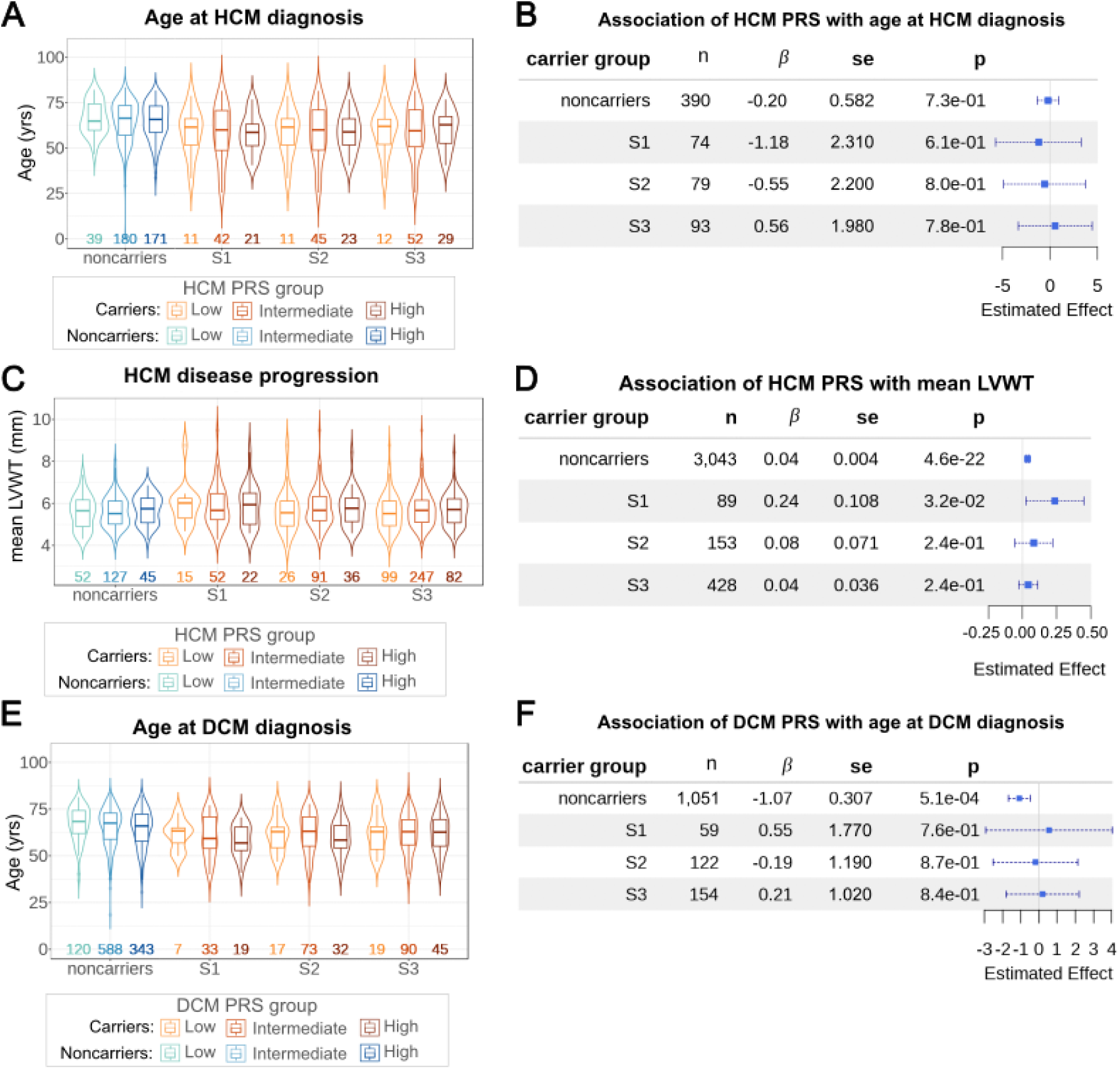
Association between HCM and DCM PRS and age of diagnosis and cardiac phenotypes among noncarriers and carriers of rare pathogenic variants. Distribution of ages of HCM diagnosis in noncarrier and carrier sets stratified by PRS bins (A). Forest plots show the association between HCM PRS and HCM age of diagnosis by linear regression among noncarrier and carrier sets (B). Distribution of mean LVWT in noncarrier and carrier sets stratified by PRS bins (C). Forest plots illustrate the association between HCM PRS and HCM mean LVWT using linear regression among noncarrier and carrier sets (D). Distribution of ages of DCM diagnosis in noncarrier and carrier sets stratified by PRS bins (E). Forest plots show the association between DCM PRS and DCM age of onset using linear regression in noncarrier and carrier sets (F). carrier sets: S1, S2, S3; PRS bins (low: bottom 20%, intermediate: middle 60%, high: top 20%); n, number of individuals in carrier sets; β, effect size; se, standard error; p, p-value.

## Supplemental Tables

Table S1. Phenotypes in the UK Biobank used in the study.

Table S2. Clinical characteristics of the study cohorts. Age at disease diagnosis, age at source censor, age at imaging assessment, body mass index (BMI) at initial assessment, and BMI at the imaging assessment are summarized by the number (n), mean, standard deviation(sd), median, and interquartile range (IQR) of the measurements. Number of individuals that are males/females, ever smoked, and have diagnosis codes for diabetes, hypertension, and hypercholesterolemia are also recorded.

Table S3. Significant variants (FDR <1%) from the GWAS of sarcomere negative HCM cases (Tadros et al 2023) used to calculate HCM PRS. SNP, rsID by dbSNP as of version 151; chr, chromosome; position, position in hg19 coordinates; gene, gene(s) overlapping lead SNP; EA, effect allele; n, estimated effective sample size; EAF, frequency of the effect allele in the meta-analysis; ßJ, effect size of the joint analysis; ßJ_se, standard error of ßJ; pJ, p-value of the joint analysis; misCount, number of subjects with missing genotype (out of 408,246 observed); misRate, rate of missing genotype data; HWE_p, Hardy-Weinberg equilibrium p-value of the variant in the UKBB participants included for PRS calculation.

Table S4. Significant variants (P-value < 5e-8) from the GWAS of LVEF (Pirruccello et al 2020) used to calculate DCM PRS. SNP, rsID by dbSNP as of version 151, chr, chromosome; position, position in Hg19 coordinates; gene, gene within 500kb of the lead SNP with the lowest GWAS P value at the locus (if any); EA, effect allele; EAF, frequency of the effect allele in the meta-analysis; ß, effect size of the GWAS analysis; p, p-value of the GWAS analysis; misCount, number of missing genotype (out of 375,804 observed); obsCount, number of observed genotype data; misRate, rate of missing genotype data; HWE_p, Hardy-Weinberg equilibrium p-value of the variant in the UKBB participants included for PRS calculation

Table S5. Schoenfeld test results to confirm the proportional hazards assumption for CoxPH models testing the association between the rare pathogenic variant classes and disease risk. PLP, pathogenic or likely pathogenic, pLoF, predicted loss-of-function; pDM, predicted damaging missense variants

Table S6. CoxPH test results with a time-varying component for the association between rare pathogenic variant classes and disease risk over time. We report the HR at birth, 25 years, 50 years, and 100 years. tt, time-varying component; β, effect size; HR, Hazards Ratio; SE, standard error; LB, lower bound; UB, upper bound. PLP, pathogenic or likely pathogenic, pLoF, predicted loss-of-function; pDM, predicted damaging missense variants

Table S7. Schoenfeld test results to confirm the proportional hazards assumption for CoxPH models testing the association between PRS and disease risk among carrier sets. S1, carriers of PLP variants; S2, carriers of PLP or pLoF variant; S3, carriers of PLP, pLoF, or pDM variants

Table S8. Association between cardiac measurements and disease status adjusted for covariates in a linear regression model.

Table S9. Number of known and predicted rare pathogenic variants within each HCM and DCM gene. Note that some carriers have multiple rare pathogenic variants.

Table S10. Number of homozygous and heterozygous carriers with rare pathogenic variants in HCM and DCM genes as identified as P/LP in ClinVar, pLoF, or pDM.

Table S11. Number of carriers with multiple rare pathogenic variants in HCM and DCM genes. Listed is the number of carriers with multiple known (likely) pathogenic variants (multi-PLP), multiple VUS/unclassified pLoF variants (multi-pLoF), multiple VUS/unclassified pDM variants (multi-pDM), known (likely) pathogenic and pLoF variants (PLP-pLoF), known (likely) pathogenic and pDM variants (PLP-pDM), pLoF and pDM variants (pLoF-pDM), known (likely) pathogenic and pLoF and pDM variants (PLP-pLoF-pDM), multiple known/predicted pathogenic variants in the same gene (multi-samegene), and known/predicted pathogenic variants in different genes (diffgenes).

Table S12. List of the known and predicted HCM and DCM variants. ClinVar Columns include: CHROMOSOME:POSITION:REF:ALT (Variant), variant ID (rsID), gene name (Gene), the gene association levels with HCM (HCM gene strength) and DCM (DCM gene strength), gene transcript (Transcript), aggregate germline CLinVar classification for this single variant (ClinVar CLNSIG), ClinVar conflicting germline classification for this single variant (ClinVar CLNSIGCONF), ClinVar review status of germline classification (ClinVar CLNREVSTAT), alternate allele count (AC), allele number (AN), alternate allele frequency (AF) within the UK Biobank exome sequence data, variant consequence based on VEP (VEP consequence), LOFTEE pLoF annotation (LOFTEE LoF), predicted deleterious missense score (AlphaMissense score), (likely) pathogenic in ClinVar (Causal Evidence), predicted high-confidence pLoF (HC pLoF), predicted deleterious missense (pDM).

Table S13. Summary of mean LVWT (mm) measurements in HCM cases, HCM controls, rare pathogenic variant carrier sets. Carriers included variants sets in HCM genes with either known evidence of being pathogenic or likely pathogenic (PLP), predicted loss-of-function (pLoF), and predicted damaging missense (pDM) variants. n, number of measurements; IQR, interquartile range.

Table S14. Summary of LVEF (%) measurements in DCM cases, DCM controls, rare pathogenic variant carrier sets. Carriers included variants sets in DCM genes with either known evidence of being pathogenic or likely pathogenic (PLP), predicted loss-of-function (pLoF), and predicted damaging missense (pDM) variants. n, number of measurements; IQR, interquartile range.

Table S15. CoxPH test results of the interaction between rare pathogenic variants and common modifiers in their effect on disease risk. HR, Hazards Ratio; SE, standard error

